# Extreme Climate Related Disasters: two-time points evaluation of the impact in children and youth mental health

**DOI:** 10.1101/2020.03.26.20044560

**Authors:** Sabrina de Sousa Magalhães, Leandro Fernandes Malloy-Diniz, Daniela Valadão Rosa, Antônio Alvim-Soares, Débora Marques de Miranda, Marco Aurélio Romano-Silva

## Abstract

Worldwide, floods and drought are the most frequent extreme climate-related disasters with a potential that might affect children and adolescent mental health. This study aimed to describe mental health impact on youth exposed to flood or drought (time 1), to provide a prospective follow up of symptoms (time 2), about 15 months apart, and to compare the child response with control groups based on ranges of socioeconomic status. Posttraumatic stress symptoms (PTSS) and general behavior problems were evaluated. Sociodemographic data from 275 children and adolescents (6 to 18 years old) were described and analyzed and they were gathered in four groups Control group with higher socioeconomic status, Control group with lower socioeconomic status, Flood group, and Drought group. At time point 1, children from all groups did not substantially differ in general behavioral problems, but PTSS scores significantly differentiate the groups. At time point 2, the Flood group presented a pattern of recovery about PTSS. For the Drought group, a persistence and stability pattern of PTSS was verified. Post-disaster longitudinal studies are essential to elucidate how psychological distress progress over time and to understand the relationship between mental health and exposure to trauma.

## 1- Introduction

Every day, a natural or technological disaster occurs on the globe [1]. In recent years, the majority of worldwide natural disasters reported were either floods and or landslides [2,3]. However, among natural disasters, drought events affected the large number of people [3]. There is medium confidence that both floods and droughts will increase in some areas [4], being the most common and critical disasters [5] in developing countries. Evidence shows that psychological consequences associated with these extreme weather events exceed physical injury by 40:1 [6]. Although the incidence of weather events have increased [6], work conducted to access the impact of climate change associated events on mental health [7] are still scarce.

Worldwide, children are estimated to bear almost 90% of disorders due to climate change [8]. Epidemiological estimates indicate that 25% of adolescents had faced a natural disaster in their lifetimes [1]. Children and adolescents presented a higher risk than adults to develop psychological distress after a disaster [9,10] due to physiological and cognitive immaturity, limited physical skills, higher metabolic rate, and dependence on others for care, protection, safety and provision [11-13]. Longitudinal studies evidenced a peak of symptoms one year after the disaster and generally an improvement over time [9,14].

After an acute-onset traumatic experience, five trajectories are commonly described: i) stress resistance in which no alteration was observed; ii) disturbance with recovery; iii) posttraumatic growth characterized by an improvement using behavioral strategies; iv) breakdown without recovery; and v) delayed breakdown without recovery [15]. The first three patterns show forms of resilience, and the others indicate maladaptive pathways. In this case, resilience was considered the capacity of positive adaptation in the face of adversity and stressors that threaten the stability, viability, or development of a dynamic system [15,16]. Verifying the presence of high levels of anxiety in the early post-disaster months, helps to predict and differentiate which children will present chronic distress or recovery pattern [17].

This study aimed to describe mental health impact on children and adolescents exposed to flood or drought, with a prospective follow up of post-traumatic symptoms, after a time lapse of about 15 months.

## 2. Method

### 2.1 Participants

Our sample was composed of 275 participants, subjects were compared according to its condition (FG/DG/Control), age (children/adolescent) and socioeconomic status (SES). Subjects 6 to 11 years old belonged to the Children subgroup, and subjects 12 to 18 years old belonged to the Adolescent subgroup. A control group was formed by 130 students recruited from three public schools at the cities of Belo Horizonte and Paraopeba, located in the southeast of Brazil. Within groups, we have made a division into higher SES e lower SES (median value of SES was used to subdivide the groups), since extreme climate events can cause a transient change in socioeconomic status. Thus, the control groups were: higher SES (HSES), children (n=42) and adolescents (n=28); lower SES (LSES), children (n=37), and adolescents (n=23).

The Flood group (FG) was formed by 61 children and 23 adolescents (n=84) from Rio Branco city, located at the northwest of Brazil. In 2015, this city faced the worst flood incident of its history. According to local public help agencies, the Acre river rose 18,4 meters, affecting 87,000 people (21% of the total population) and leaving 10,400 unsheltered after the incident. Time 1 survey was conducted 40 days after the peak of the disaster. Subjects were recruited from a local public school and social services family users. Drought group (DG) was composed of 27 children and 34 adolescents (n=61) from Francisco Sá, located at the southeast of Brazil, and belongs to semi-arid zone, with total annual pluviometric indices inferior to 1.000 mm [18]. They were recruited from a rural and an urban public school. The two main features for the last groups were: facing a natural hazard (flood or drought) and social disadvantages.

We did not collect data from control groups at the follow-up. Demographic characteristics of the sample are shown in tables 1 and 2, respectively, for children and adolescents. We tried to match groups according to age, gender, and SES. The HSES group provided a measure of expected child development in the absence of deprivation in the same country.

**Table 1.a).** Demographic characterization of the study children sample, and data about the equivalence between groups according to age and gender (n=167)

|  | Children | Gender (F/M) | Age - M (SD) | SES |
| --- | --- | --- | --- | --- |
| **HSES (I)** | n=42 | 25/17 | 8,74 (1,61) | 33,60 (10,46) |
| **LSES (II)** | n=37 | 16/21 | 8,81 (1,65) | 16,46 (2,88) |
| **Flood group (III)** | n=61 | 30/31 | 8,57 (1,30) | 17,74 (5,43) |
| **Drought group (IV)** | n=27 | 15/12 | 8,62 (1,83) | 12,52 (8,16)<sup>a</sup> |
| **Statistics** | | $\chi^2 = 2,402$,<br>$p=0,493$ | F(3,167) =<br>0,786, $p=0,853$ | **F(3,161) =**<br>**88,497, $p < 0,001$** |
| **Pos hoc** | | - | - | HSES is different<br>from all groups;<br>and III ≠ IV |

Note: HSES: Control group with higher socioeconomic status; LSES: Control group with lower socioeconomic status; F/M: female/male; M: mean; SD: standard deviation; SES: socioeconomic status. <sup>a</sup>n=21.

**Table 1.b).** Demographic characterization of the adolescent sample, and data on the equivalence between groups according to gender (n=108).

|  | Adolescents | Gender (F/M) | Age - M (SD) | SES |
| --- | --- | --- | --- | --- |
| **HSES (I)** | n=28 | 22/6 | 13,61 (0,99) | 31,32 (6,97) |
| **LSES (II)** | n=23 | 11/12 | 13,00 (1,00) | 15,78 (3,22) |
| **Flood group (III)** | n=23 | 11/12 | 14,52 (2,13) | 13,91 (4,41) |
| **Drought group (IV)** | n=34 | 19/15 | 14,20 (1,61) | 21,64 (7,49)<sup>a</sup> |
| **Statistics** | | $\chi^2 = 6,892$,<br>$p=0,075$ | **F (3,107) = 16,589**,<br>**$p=0,001$** | **F (3,99) = 58,693**,<br>**$p < 0,001$** |
| **Pos hoc** | | - | II≠III; II≠IV | HSES is different<br>from all groups;<br>and III≠IV |

Note: HSES: Control group with higher socioeconomic status; LSES: Control group with lower socioeconomic status; F/M: female/male; M: mean; SD: standard deviation; SES: socioeconomic status. <sup>a</sup>n=25.

**Table 2.a).** Posttraumatic Stress Symptoms in children, according to CRIES, and differences between groups in time 1.

| CRIES | HSES (I) | LSES (II) | Flood group (III) | Drought group (IV) | Statistics | Pos hoc | Effect size ($d_{\text{Cohen}}$) |
| --- | --- | --- | --- | --- | --- | --- | --- |
|  | Children (n=42) | Children (n=37) | Children (n=54) | Children (n=25) |  |  |  |
| Intrusion | 3,62 (4,87) | 2,41 (3,77) | 7,28 (5,42) | 3,56 (4,44) | $\chi^2(3)=26,150, p<0,001$ | I≠III; II≠III; III≠IV | 0,84 |
| Avoidance | 4,26 (5,34) | 3,22 (4,08) | 10,37 (5,19) | 4,04 (4,95) | $\chi^2(3)=46,190, p<0,001$ | I≠III; II≠III; III≠IV | 1,25 |
| Total | 7,88 (9,42) | 5,62 (7,17) | 17,65 (9,27) | 7,60 (8,47) | $\chi^2(3)=40,824, p<0,001$ | I≠III; II≠III; III≠IV | 1,14 |

Note: HSES: Control group with higher socioeconomic status; LSES: Control group with lower socioeconomic status.

**Table 2.b).** Posttraumatic Stress Symptoms in adolescents, according to CRIES, and differences between groups in time 1.

| CRIES | HSES (I) | LSES (II) | Flood group (III) | Drought group (IV) | Statistics | Pos hoc | Effect size ($d_{\text{Cohen}}$) |
| --- | --- | --- | --- | --- | --- | --- | --- |
|  | Adolescents (n=28) | Adolescents (n=22) | Adolescents (n=19) | Adolescents (n=34) |  |  |  |
| Intrusion | 4,57 (6,14) | 3,18 (4,64) | 11,74 (4,29) | 6,88 (5,12) | $\chi^2(3)=28,987, p<0,001$ | I≠III; II≠III; II≠IV; III≠IV | 1,19 |
| Avoidance | 4,14 (5,65) | 3,86 (5,99) | 12,11 (4,53) | 5,53 (4,72) | $\chi^2(3)=26,806, p<0,001$ | I≠III; II≠III; III≠IV | 1,13 |
| Total | 8,71 (10,71) | 7,05 (10,49) | 23,84 (6,78) | 12,41 (7,92) | $\chi^2(3)=32,228, p<0,001$ | I≠III; II≠III; III≠IV | 1,29 |

Note: HSES: Control group with higher socioeconomic status; LSES: Control group with lower socioeconomic status.

Inclusion criteria were written informed consent by parent or a proper caregiver, minimum age of six years old. Exclusion criteria were any current or previous diagnosis of epilepsy or any neurological disorder. A total of 17 children (5,5% of the total sample) presented some health issue informed by parents, regardless of the group.

### 2.2 Assessment

#### 2.2.1 CRIES

Children’s Revised Impact Scale (CRIES-8) is a screening tool for PTSS which evaluate the presence of symptoms during last week, in children and adolescences. It consists of eight items, half of them measures intrusion complaints associated with the stressful event, and the other half measures avoidance behavior related to the event. Intrusion and Avoidance subscales are obtained through the sum of the individual items [19]. Details about the scoring point of the scale are described elsewhere [19].

#### 2.2.2 CBCL

Child Behavior Checklist (CBCL) consists of screening questions about behavioral problems in school-aged children, from 6 to 18 years old, answered by the parent or caregiver. CBCL provides subscales grouped according to a superordinate category: I) internalizing problems: anxious/depressed, withdrawn-depressed, somatic complaints, social problems, thought problems, and attention problems; II) externalizing problems: rule-breaking behavior and aggressive behavior; III) total problems; IV) subscales based on DSM-IV: affective problems, anxiety problems, somatic problems, attention deficit/hyperactivity (ADH) problems, oppositional defiant problems, and conduct problems. Answers were transformed to *t* scores to indicate the performance of the individual in reference to normative data concerning culture (i.e., Brazilian), age, and gender [20].

#### 2.2.3 CCEB

Brazil’s Economic Classification Criteria (CCEB) is a tool to access and classify SES through investigation of some domestic features, like the existence and quantity of some domestic items and the years of education of family’s representative member [21].

### 2.3 Statistical analysis

Statistical Package for the Social Sciences 20.0 (SPSS) platform was used to perform statistical analysis and graphs construction. Descriptive analysis indicated the demographic profile of the sample and performance on mental health scales. The vast majority of data presented a normal distribution. However, we decided to adopt more rigorous criteria, and nonparametric tests (Kruskal-Wallis Test) were used to verify the difference among the groups.

### 2.4 Procedures

After formal contact with schools or social service, parents were invited to engage in our study. Both parents and children read the term of participation and adults gave written informed consent and youth signed the assent term.

Local ethics committee approved all procedures of this study under the registration number CAAE: 26886814.9.0000.5149. We interviewed parents and kids once each and separately. Both interviews occurred in one meeting that lasted around 90 minutes. At prospective follow-up, we re-evaluated children’s mental health. Follow-up of the flood condition was after 14 months, and of the drought condition after 17 months. The control groups did not have a second evaluation.

## 3. Results

### 3.1 Disaster experience

All family participants in FG declared to be affected by 2015’s flood. For more than 80% of this group, a flood incident had already occurred before. Youth reported feeling scared (73%), and they believe to be in risk of death (79%). Families from DG had water supplied by general public services with a water rationing program that provided water day-on day-off for the community, although 72% of families declared being affected by the drought. Concerns about drought and scarcity of water were reported for almost our entire youth sample, as well as the feeling of danger due to water scarcity. More than half affirmed that the actual drought conditions interfered with their future plans. Mostly of the Control group did not have previous experience of climate events and did not report any significant stressful life experience. Demographic data is in Table 1a and 1b.

### 3.2 Mental health at time point 1

CRIES scores showed significant differences related to PTSS. Table 2a displays data from children and Table 2b from adolescents. Children and adolescents from the Flood group presented the higher scores for intrusion and avoidance behavior related to the natural hazard and total score of CRIES. Youth from DG did not significantly differ from the control groups. If we considered a cut-off score of 17 in CRIES^29^ as a positive screening for PTSD, we found a prevalence of 57,4% of children from FG; 94,7% of adolescents from FG; 16,0% of children from DG; 23,5% of adolescents from DG.

To better characterize the experience of disaster, we analyzed qualitative parameters and CRIES outcomes. For both natural hazards groups, feeling of being in danger did not correlate with any CRIES measures. On the other hand, children from DG concerns about their own future significantly correlated with the total score of CRIES (r=0,41, p<0,05). In the adolescents group, this distress correlated with the CRIES avoidance score (r=0,37, p<0,05).

The evaluation of symptoms of psychiatric domains using CBCL did not indicate major differences between groups. For children, significant differences, with medium effect sizes, were achieved for the domain of affective problems between the disasters groups (FG and DG), somatic problems differences were more reported for FG, and symptoms of attention and hyperactivity deficit were prevalent for LSES group of children. For adolescents, CBCL presents only differences in Anxious/Depressed domain, but between controls since the LSES group report more symptoms in this domain than HSES. There is none difference in reports from DG or FG compared to Controls.

### 3.3 Mental health at time point 2

After 14 months for FG, and 17 months for DG, we conducted a prospective follow-up. Children responded to CRIES according to the natural hazard they were exposed. In FG, children’s CRIES scores did not statistically change over time. For adolescents, we found a significant decrease in PTSS symptoms with large effect sizes (see Figure 2). Considering a cut-off value of 17 [22], the prevalence of PTSD at time point 2 was 45,6% of FG’s children; 36,3% of FG’s adolescents; 23,8% of DG’s children; and 39,1% of DG’s adolescents.

**Figure 1.**
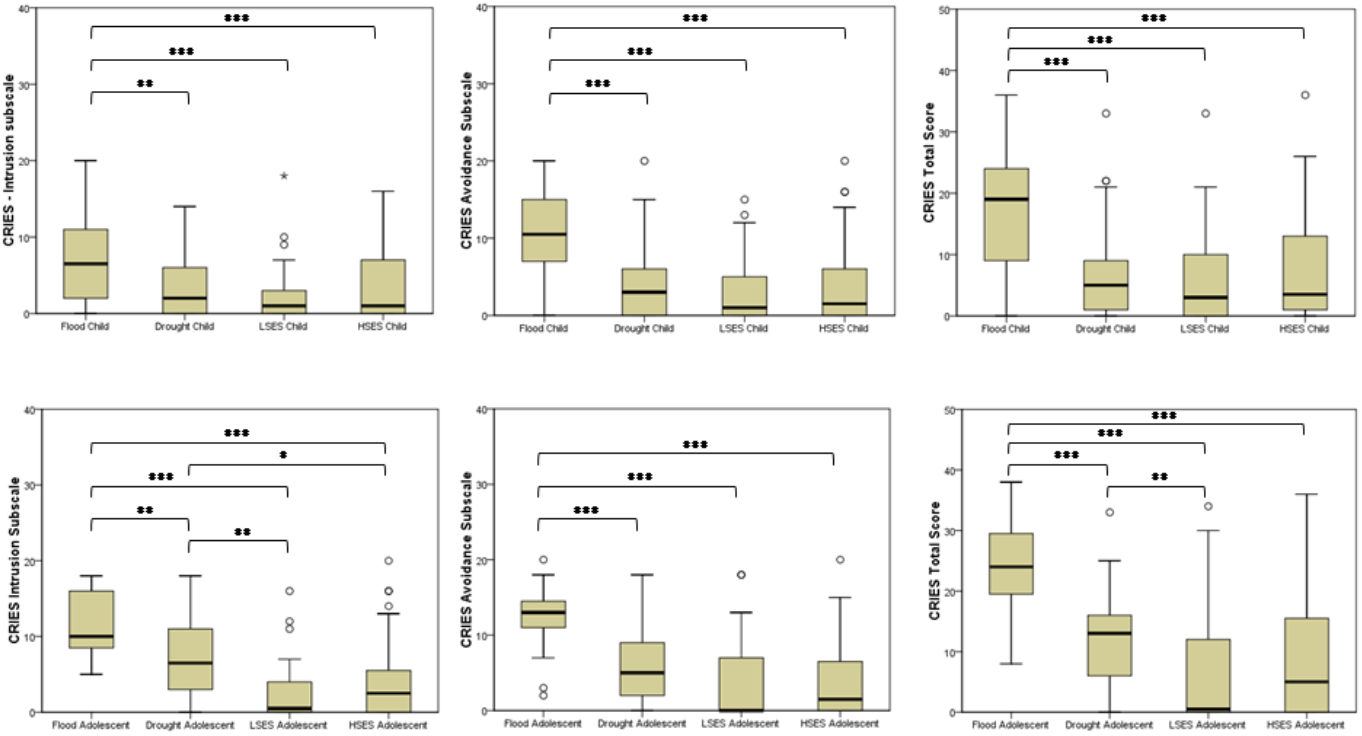
Comparative differences in CRIES scores between groups. Upper panel exhibits data for children subjects, and lower panel for adolescents participants. (*Note*: SD: social disadvantage, \**p* < 0,05, \*\**p*< 0,01, \*\*\**p*<0,001).

**Figure 2.**
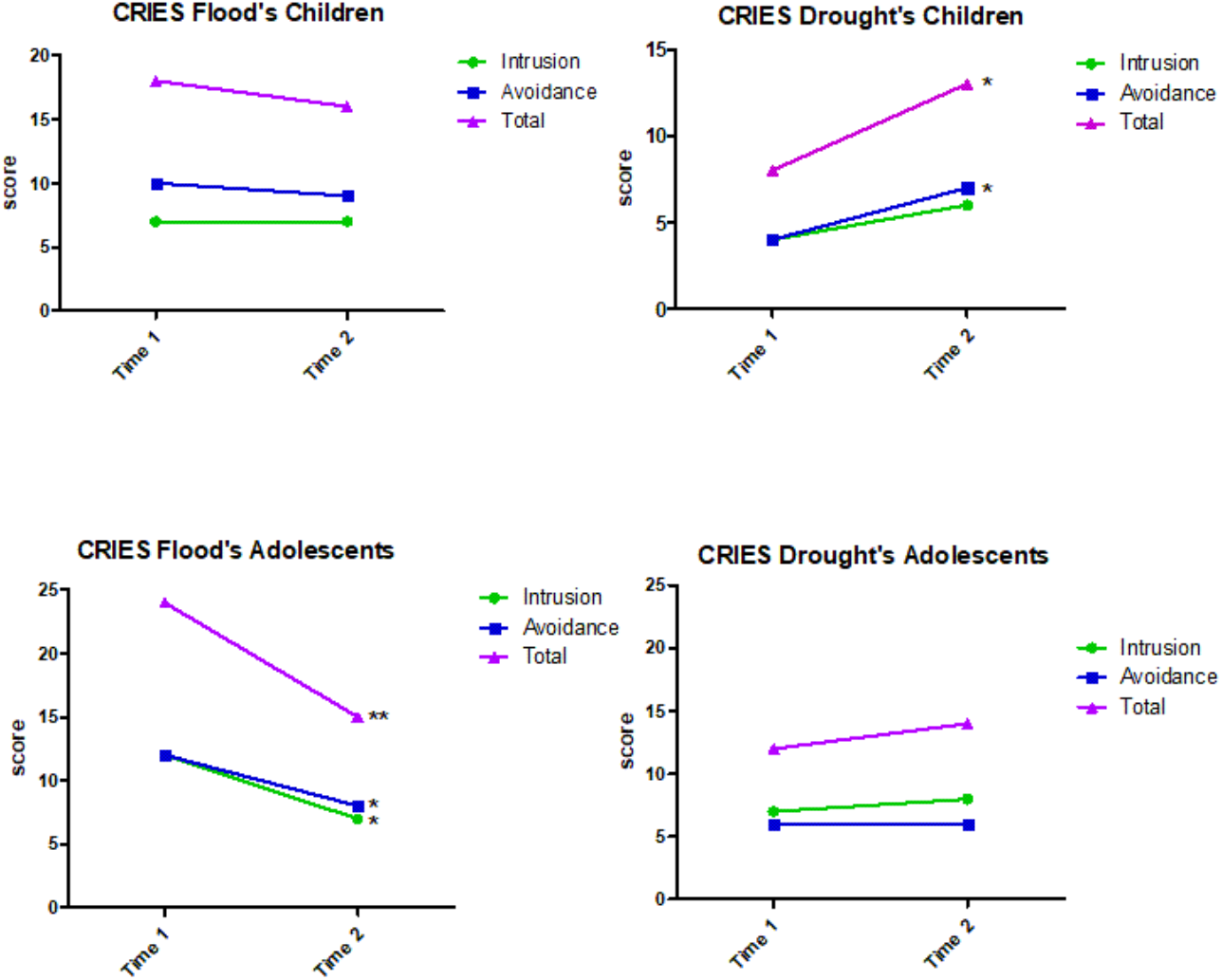
CRIES scores between the two time points evaluations for both natural hazards groups and age groups. (*Note*: * *p* < 0,05, \*\**p*< 0,01)

Follow up results for DG indicated that, for PTSS, we found stability of complaints for DG adolescents, and a significant increased pattern of complaints in the avoidance domain and total score of CRIES for children. However, none of the mean scores of groups reached the cut-off of 17 points.

## 4. Discussion

We addressed mental health impact of climate change-related disasters on children and adolescents through evaluation of general behavior problems and PTSS on subjects exposed either to flood or drought conditions. We also followed-up the trajectories of PTSS over a year after the first evaluation. All climate events happened in Brazil in conditions very similar to disasters in low or middle-income countries [2].

Youth is a vulnerable group to suffer adverse effects of extreme weather events. The rationale includes their initial developing coping capacity, limited capacity to mobilize resources to assist them independently, a higher dependency of a caregiver, developmental timing and sensitiveness [23]. All children from FG feels as direct or indirectly affected by the 2015 flood, and the vast majority of them had been previously exposed to such incident. While not all families from DG reported being affected by droughts, we observed indirect impact in all of them, since water distribution by public agency has already been rationed during all year due to scarcity. Howe et al. [24] described the need of at least 25 weeks of severe drought conditions in order to the majority of people report experiencing a drought event. Drought disaster and its impacts are so common that it is challenging to reach proper allocation of needed resources, and community engagement in preventive and mitigation actions.

### 4.1 Mental health at time point 1

Youth just exposed to disasters present a higher risk for developing an extensive set of responses, including PTSS, depression, anxiety, sleep disorders, attachment disorders, aggression, functional impairment, substance use, suicidal ideation and behaviors, risk-taking behaviors or other mental health disturbances [10,, 25-27]. In consequence, a cascade of interference may disrupt children’s emotion regulation, behavioral control, cognition, learning, language development, and academic performance, which in turn can influence adverse mental health outcomes [10,14]. The prevalence of mental health symptoms in children exposed to disaster can be 44 to 104% superior to pre-disaster baseline, up to two years after the incident [28].

At point 1, children from the natural hazards groups exhibited minor behavioral problems with medium effect size. Few studies did not find any youth mental health consequences after experiencing a disaster [29]. Only affective and somatic problems domain were more frequently reported in FG children. These symptoms could be highlighted due to change in routine and the immersion in stressful condition. One hypothesis is a possible change in parents and caregivers perceptions about their children [30,31]. Parent monitoring towards children after a disaster can raise, be exacerbated in an unreal way, decreased, or even be distant in the sense that they do not express concern about the possible psychological consequences for children [30-33]. Another hypothesis is based on the influence of resilient behavior. After all, adverse experiences are not necessarily linked with adverse psychological responses. Most children exhibit minimal symptoms following disasters [17,34] or even do not develop any psychopathology, exhibiting a continued capacity of functioning [35]. Children can exhibit more resilience than adults and recover faster in some circumstances [13]. Resilience does not implicate absence of any initial psychological distress; instead, it refers to the ability of “bouncing back” [35]. A child who exhibits resilience may show better personal resources if faced again with a future traumatic event [36].

As a sub-acute or chronic hazard, drought may impact youth to a greater extent in indirectly ways [6], undetectable by screening tests like CBCL. Vins et al. [37] indicated that the primary pathway connecting drought and mental health was via economic impact of land degradation. Consequences in economic structure (i.e. availability of job offers, income insecurity, and disruption of physical and social infrastructure) influence mental health in long term or with earlier subthreshold symptoms. Adolescents from the drought group had increased anxiety symptoms, and also perceived their future and financial situation as threatened by the possibility of aggravation of the drought scenario. Droughts impose some constraints on employability in their hometown, the need to help family income, sometimes giving up plans or imposing the need of migration [38]. Correlation between fear of the future and CRIES’ avoidance score implies a tendency to cognitive avoidance and an attempt to avert the dealing with internal distress that the current context already provoked on them.

As the preponderant response after an extreme event [35], PTSS scores differentiate groups and CRIES scores provided some interesting insights about youth reaction in response to the disaster. Ultimately, PTSD outcome in children and adolescents is mostly a reaction to the specificities of the event, instead of previous functioning or experience [39]. FG presented higher rates of intrusion and avoidance symptoms, and also higher total CRIES score. Flood incidents seems to be more disturbing and to elicit more post-event PTSS than droughts. Flood is considered an acute hazard and is more related to direct mental health consequences [6].

Prevalence of PTSD varies significantly among studies [12,40], ranging from 5% to 30% [10,40] but can reach even 60,8% of exposed youth [41]. Prevalence rate of probable PTSD in our study was significantly higher for FG, compared to the DG, and both controls groups. Chen et al. [42] evaluated PTSS on Chinese youth (aged 8-18 years) exposed to a flood event, using CRIES. They verified that 46,6% of participants presented probable PTSD, and older children exhibited higher values on total CRIES score. Our results from the FG were compatible with those described for Chinese children. Evaluating emotional distress using CRIES, made it possible to reliably detect subthreshold cases that are very important to recognize in order to provide early specialized intervention [43]. So, it is crucial to mitigate the impact of any hazard and avoid new stressful events.

### 4.2 Mental health at time point 2

Prospective follow-ups of a disaster are still rare. However, they are essential to identify the course of PTSD in order to differentiate pathological and normative responses, onset, and remission [44]. After a natural disaster, the prevalence of PTSD and PTSS can range from 14,0% three months post-event to 30,6% in children one year after [45,46]. Nevertheless, chronic PTSS rarely increase to over 30% of the sample [34]. Yet years after flood or drought incidences, symptoms of PTSD, emotional distress behavioral difficulties, and disturbance in family functioning and community dynamics can be still observed [47,48]. One important asset for the recovery from prior PTSD is social support.

After more than one year after assessment of PTSS, FG children did not exhibit symptom remission over time, although showed a tendency for a decrease. In adolescents, we found a substantial decrease of symptoms, with mean score below the cut-off for PTSD screening [22]. At follow-up, children from the DG presented higher scores of PTSS, especially at the avoidance domain, although for adolescents no change was observed. Prevalence of positive screening for PTSD at time point 2 was still higher for children from FG and adolescents from DG.

For FG, a typical acute-onset event, PTSS trajectories are distinct for children and adolescents. Younger individuals exhibit disturbance in the first assessment, and a tendency to recover in following evaluations, an indicative of resilient behavior. Adolescents presented a marked remission of PTSS, thus denoting recovery and resilience [15,35]. In the drought scenario, we could not apply the pathways to chronic-onset extreme event indicated by Masten and Narayan [15] as the adverse effects were still ongoing with lack of restored conditions. However, we did observe that children with previous normal functions showed a tendency to a slight breakdown with the increase of persistent PTSS; thus, it could be characterized as a resistance pattern [35]. For adolescents, PTSS remained stable, below cut-off, but slightly increased in the second evaluation.

Flood is a typical rapid-onset, short-duration extreme event, tending to be short-lived, and is usually associated with short-term PTSD symptoms; whereas drought, a slower-onset and chronic disaster that accumulate adversities and vulnerabilities, lead to long term severe mental health outcomes [10]. Noteworthy is the intensification of avoidance complaints in children from DG. Trickey et al. [39] validated thought suppression as one of the main risk factors to youth PTSD, and they concluded that the avoidance cluster was central to PTSD symptomatology. In the time considered in this study, children did not reach the threshold for positive PTSD. However, avoidance scores correlated with fear about the future in our sample.

An open-ended question on the field is about the effects of repeated disasters on an individual’s mental health. In the Northern city, flood events are unfortunately recurrent every rainy season, and, in turn, drought events are a chronic condition in the semi-arid zone of the Brazilian Northeast region. Previous exposure to extreme adversity can change the response to a subsequent traumatic event [15]. Our data suggested a protective effect (stability or remission of symptoms) on children exposed to recurrent flood, and a sensitizing model for drought youth. A remission pattern was previously reported [49-51], as resilience and emotional coping can explain up to 30% of the variance for PTSS, in a model validated with adolescents exposed to an earthquake [52]. To count with a resilience repertoire does not mean that no adverse symptom will emerge. On the contrary, stress level can interfere in resilience behaviors and lead to manifestations of PTSD, depression, or other subclinical psychiatric symptoms [53,54]. Resilience repertoire will become visible as the ability to bounce back to a previous healthier state and regain prior functionality [35].

In drought context, the prospect is different. Youth from a drought scenario are continuously exposed and live within the secondary adversities imposed by drought. We verified stability or enhancement of symptoms. Stability and persistence of complaints were described by Thienkrua et al. [55], Osofsky et al. [56] and Jia et al. [57]. A vulnerability-inducing effect, or sensitizing model, can account for this pattern of finding and the maintenance of problems related to drought effects contribute to it.

Brazilian’s flooded community was already engaged in dealing with flood events, and the municipality had initiated mitigation and preventive actions. Their social preparedness for disaster can partially explain the findings of mental health symptoms, as social systems that directly interact with children exposed to disaster are essential contributors to children response. Besides, resilience behaviors, positive adaptation, and protective effects have the potential to spread within individuals, families, communities, and across generations [15]. Therefore, to ensure familiar and social stability, and to strengthen these systems are a meaningful way to assist children after disasters and to confer resilience [15, 58-60].

Interventions delivered in the post-disaster period may attenuate short-term PTSS and impairment. Special attention must be addressed to anxiety complaints in the months immediately post-disaster, as they are linked to future outcomes [17]. Alleviating secondary stressors may be crucial in dealing with the long-term course of posttraumatic stress and in restoring functionality and quality of life [61].

### 4.4 Limitations

Methodological challenges in context of disaster studies impose some constraints that may influence results and limit generalizability of findings [35]. Both our disaster samples were very heterogeneous and included youth directly and indirectly affected by the extreme event, which *per se* may lead to lower estimates of the psychological burden. Further, we rely mostly on a convenience sample, and representativeness of our sample can be questionable. Due to practical field conditions, we chose to collect data through school partnerships. Students from same school setting form a “cluster” that may compromise the degree of representativeness stated above [62].

The prospective methodology supports better the understanding of disaster influence on youth mental health and trajectories of symptoms. Unfortunately, we lost some participants due to migration or impossibility to contact during the second assessment wave. Comparison with non-affected communities provides useful insights, but some crucial differences between groups cannot be suppressed. Thus, our methodological choices did not imply causality. Lack of pre-disaster information is also a limitation.

### 4.5 Conclusion

Extreme climate events are stressful circumstances with different after-disaster findings in children and adolescents, with long-term consequences of childhood trauma [43]. Public health systems must be prepared to deal with children’s reaction to natural hazards, as the odds of an adverse outcome are high. In addition, the number of children exposed to disaster is prone to increase each year, and the occurrence of an extreme event may alter community and geographic vulnerability to future events [4]. Previous social determinants interact with the characteristics of the disaster itself, imposing a challenge to disentangle both influences [6].

This study fulfills the required agenda of research in the field of mental health and climate changes [7]. Furthermore, youth population answer to a disaster is understudied. Our work provides valuable insights for mitigation strategies and the development of preparedness actions. For developing countries, focus should be on youth whose vulnerability to the effects of climate change is higher than any other [10].

Children from DG presented an intensification of PTSS and adolescents exhibited a stabilization of symptoms after more than one year. Chronicity, SES, and poverty may have contributed to outcome data, and its influence cannot be neglected. On the other hand, children from FG did not alter the patter of PTSS presented as opposed to the adolescents which significantly decreased the PTSS complains.

Longitudinal studies beyond one-year post-disaster are unfortunately rare; however, they are critical to elucidate how original patterns of psychological distress progress over time [34]. Just a few individuals will need specialized mental healthcare to meet their psychosocial needs [63], however, it is critical to accurately identify any mental disorder symptoms from disaster-related emotional distress. Allocation of resources, development of public policies, treatment options, and adapted interventions rely on that outcome. Increasingly recognition of human capacity to adapt to stressor challenges the simplistic vision that a stressful event undoubtedly results in psychopathological outcomes. Our data provided empirical evidence for the resilience capacity of children to cope well amid a disaster. Generally, symptoms remit over time, although subclinical complaints can persist.

## Data Availability

Our ethics approval does not allow data sharing.

## Acknowledgments

Financial support: CNPq 404047/2013-0 and CAPES/Brazil.

## Suplemmentary material

**Table 1.** CBCL t scores for children, according to the group, and analysis of differences between them.

| CBCL | HSES (I) | LSES (II) | Flood group (III) | Drought group (IV) | Statistics<sup>a</sup> | Pos hoc | Effect size (*d* Cohen) |
| --- | --- | --- | --- | --- | --- | --- | --- |
|  | Children (n=39) | Children (n=36) | Children (n=44) | Children (n=21) |  |  |  |
| Anxious depressed | 54,15 (11,21) | 55,83 (11,04) | 54,11 (9,86) | 52,33 (11,94) | $\chi^2 (3) = 2,188, p=0,534$ | - | - |
| Withdraw depressed | 45,44 (9,72) | 48,53 (8,18) | 47,02 (9,62) | 47,33 (11,41) | $\chi^2 (3) = 4,365, p=0,225$ | - | - |
| Somatic complaints | 47,92 (6,82) | 48,33 (8,67) | 51,45 (10,42) | 47,43 (12,25) | $\chi^2 (3) = 4,343, p=0,227$ | - | - |
| Social problems | 48,08 (7,35) | 49,53 (9,83) | 49,95 (8,20) | 46,10 (10,73) | $\chi^2 (3) = 6,309, p=0,097$ | - | - |
| Thought problems | 51,38 (11,87) | 51,89 (10,80) | 50,77 (11,07) | 52,57 (14,29) | $\chi^2 (3) = 0,676, p=0,879$ | - | - |
| Attention problems | 46,77 (7,96) | 50,58 (11,21) | 49,27 (9,89) | 47,62 (11,07) | $\chi^2 (3) = 2,468, p=0,481$ | - | - |
| Rule-breaking behavior | 45,74 (4,65) | 44,83 (4,74) | 46,68 (6,94) | 45,19 (5,66) | $\chi^2 (3) = 1,268, p=0,737$ | - | - |
| Aggressive behavior | 50,26 (9,28) | 51,86 (8,98) | 50,43 (10,51) | 49,38 (9,70) | $\chi^2 (3) = 2,095, p=0,553$ | - | - |
| Internalizing problems | 49,46 (9,23) | 51,47 (8,86) | 51,27 (8,80) | 49,05 (12,67) | $\chi^2 (3) = 4,400, p=0,221$ | - | - |
| Externalizing problems | 47,33 (8,02) | 49,28 (9,40) | 48,61 (11,41) | 46,62 (7,81) | $\chi^2 (3) = 1,362, p=0,714$ | - | - |
| Total problems | 48,41 (7,44) | 51,14 (8,55) | 50,23 (8,48) | 47,81 (12,09) | $\chi^2 (3) = 5,657, p=0,130$ | - | - |
| Affective problems | 47,08 (6,26) | 47,06 (8,11) | 48,61 (6,87) | 45,43 (12,75) | $\chi^2 (3) = 8,443, p=0,038$ | III≠IV | 0,41 (medium effect) |
| Anxiety problems | 50,41 (8,69) | 55,14 (12,93) | 49,82 (9,60) | 53,05 (12,84) | $\chi^2 (3) = 3,133, p=0,372$ | - | - |
| Somatic problems | 47,28 (6,85) | 46,47 (8,37) | 51,82 (11,52) | 47,86 (13,55) | $\chi^2 (3) = 7,931, p=0,047$ | II≠III; III≠IV. | 0,39 (medium effect) |
| ADH problems | 46,44 (7,39) | 52,25 (10,68) | 50,86 (10,27) | 47,71 (11,42) | $\chi^2 (3) = 7,971, p=0,047$ | I≠II; II≠IV | 0,39 (medium effect) |
| Oppositional defiant problems | 51,44 (10,10) | 49,17 (9,56) | 49,80 (12,08) | 49,14 (11,06) | $\chi^2 (3) = 1,623, p=0,654$ | - | - |
| Conduct problems | 46,41 (6,37) | 45,64 (5,70) | 47,80 (8,03) | 45,62 (4,26) | $\chi^2 (3) = 0,913, p=0,822$ | - | - |

*Note:* HSES: Control group with higher socioeconomic status; LSES: Control group with lower socioeconomic status; ADH: Attention Deficit Hyperactivity ; <sup>a</sup>Kruskal-Wallis analysis.

